# On mobility trends analysis of COVID-19 dissemination in Mexico City

**DOI:** 10.1101/2021.01.24.21250406

**Authors:** Kernel Prieto, M. Victoria Chávez-Hernández, Jhoana P. RomeroLeiton

**Author notes:** Email id, Email id.

## Abstract

This work presents a forecast of the spread of the new coronavirus in Mexico City based on a mathematical model with metapopulation structure by using Bayesian Statistics inspired in a data-driven approach. The mobility of humans on a daily basis in Mexico City is mathematically represented by a origin-destination matrix using the open mobility data from Google and a Transportation Mexican Survey. This matrix, is incorporated in a compartmental model. We calibrate the model against borough-level incidence data collected between February 27, 2020 and October 27, 2020 using Bayesian inference to estimate critical epidemiological characteristics associated with the coronavirus spread. Since working with metapopulation models lead to rather high computational time consume, we do a clustering analysis based on mobility trends in order to work on these clusters of borough separately instead of taken all the boroughs together at once. This clustering analysis could be implemented in smaller or lager scale in different part of the world. In addition, this clustering analysis is divided in the phases that the government of Mexico City has set up to restrict the individuals movement in the city. Also, we calculate the reproductive number in Mexico City using the next generation operator method and the inferred model parameters. The analysis of mobility trends can be helpful in public health decisions.

## 1 Introduction

Coronavirus disease 2019 (COVID-19) is an illness caused by a novel coronavirus. Coronaviruses are a family of viruses that cause infection in humans and animals. Diseases by coronavirus are zoonotic [58]. Coronaviruses that affect humans (HCoV) can produce clinical symptoms including Severe Acute Respiratory Syndrome (SARS) viruses and Middle East Respiratory Syndrome (MERS-CoV) [45]. This disease was first identified amid an outbreak of respiratory illness cases in Wuhan City, Hubei Province, China. This disease was initially reported to the WHO on December 31, 2019. On March 11, 2020, the WHO declared COVID-19 a global pandemic [32]. Since the beginning of the epidemic, to January 21, 2021 have been reported more than 97,890,676 cases and more than 2,094,459 deaths in the world.

The first case of COVID-19 in South America was registered in Brazil on February 26, 2020. The first death from this infection in this region was announced in Argentina on March 7, 2020. Then, the virus arrived to Mexico, where it is now becoming a large problem with almost 1,688,944 confirmed cases and 144,371 deaths to January 21, 2021. To date, many researchers around the world have focused their interests on understanding the transmission dynamics of COVID-19 disease using mathematical and statistical models and methods, see e.g., [16, 17, 43, 51, 53, 60, 61, 62, 63], but we will focus on those that incorporate information on human movement. A relation between human mobility and transmission of coronavirus disease in USA has been studied in [41, 46]. Metapopulation models are one of the simplest spatial models, but they are also one of the most applicable to modelling many human diseases [34]. The metapopulation concept is to subdivide the entire population into distinct *sub-populations*, each of which has independent dynamics, together with limited interaction between the sub-populations. This approach has been used to great effect within the ecological literature [33] and recently to model the spread of COVID-19 see e.g., [10, 20, 35, 38, 57].

In this work, we calibrate the metapopulation model proposed by Li et. al. in [35], similar to [64], using incidence data reported in [25]. For this end, we first describe the mathematical model used, then we computed the number of trips that are produced and attracted in some borough of Mexico City using data about these trips in 2017 [30] and combining them with the rates of reduction or increase in mobility during the pandemic reported by Google [27] and the government of Mexico City [24]. Then, by using Bayesian inference we solve the associated inverse problem to predict the dynamics of the spread of cases similarly as in the references [4, 5, 36, 40, 44, 47, 52]. Conclusions are presented in the last section.

## 2 Computation of the mobility matrices

In order to incorporate the mobility in the transmission model, the produced and attracted trips in the boroughs of Mexico City are considered (see Figure 1 and Table 1). Mexico City is the capital of Mexico, with around 9 million inhabitants and floating population of over 22 million composed of daily commuters and international visitors. Mexico city belongs to the top ten most crowded cities in the world [59]. It has a large number of headquarters and a large transport network, composed of 20 different modes of transport.

**Table 1:**
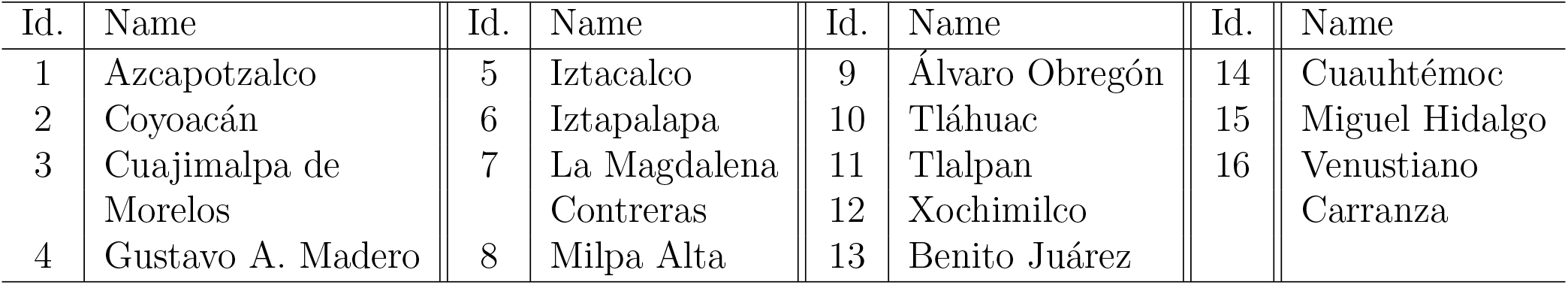
Municipalities of Mexico City.

**Figure 1:**
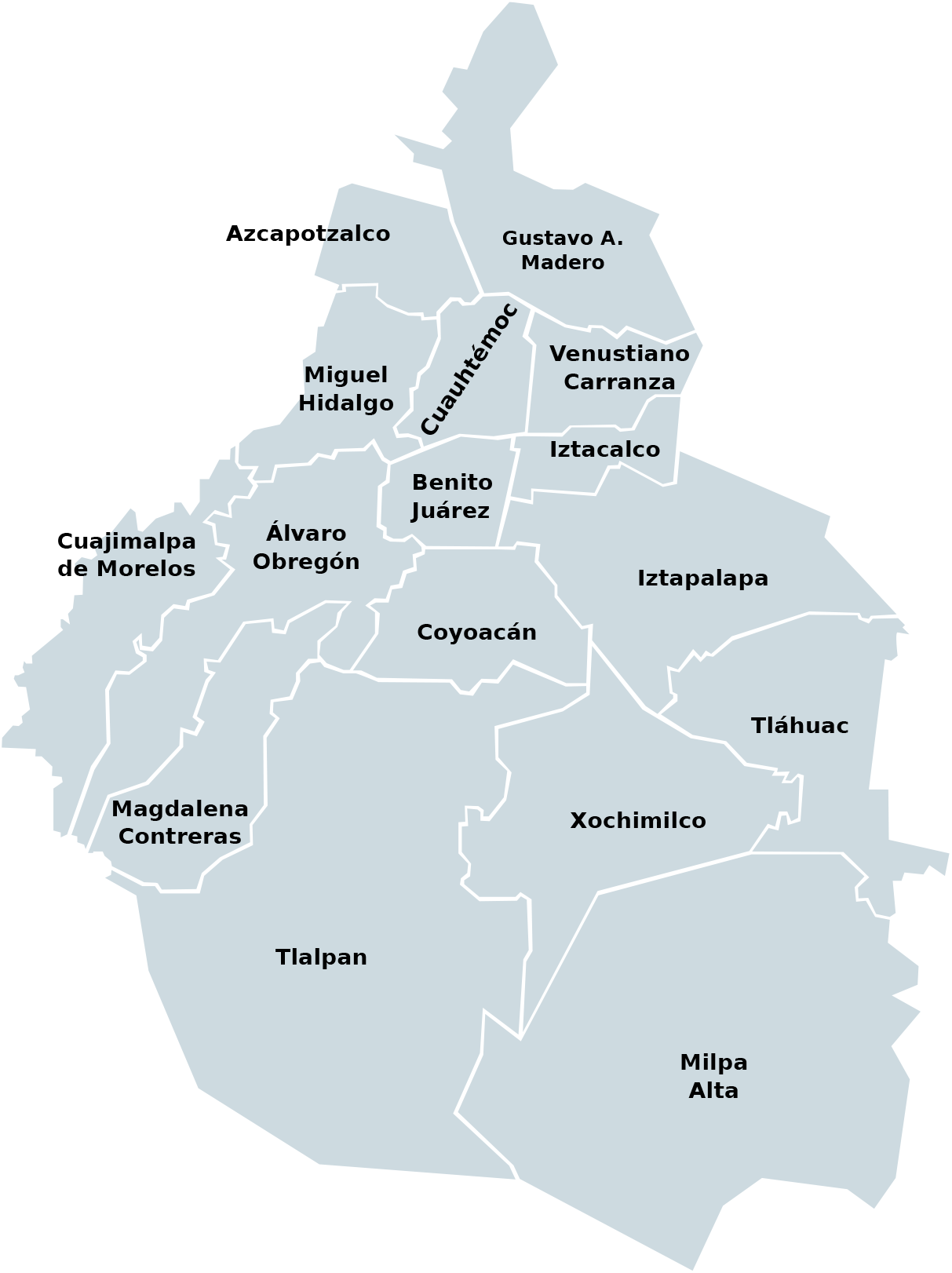
Municipalities of Mexico City.

The mobility between the zones in Table 1 is represented in a two-dimensional arrangement known as the origin-destination matrix (O-D matrix) **M** = {*M*_*ij*_}, *i, j* = 1, .., 9, where *M*_*ij*_ represents the number of trips from zone *i* to zone *j*. Origin-destination matrices are usually obtained every 10 years from surveys, in Mexico City the last one was carried out in 2017 [30]. The information available identifies, among other things, if the trip was made on a weekday or if it was made during the weekend, the transport mode used, the purpose and the time. In this paper, we consider all the trips and are identified by the mode of transport that was used to carry out in the area of interest (see Table 2).

**Table 2:** Transport modes used in the area of interest.

|  |  |  |  |  |  |  |
| --- | --- | --- | --- | --- | --- | --- |
| Car | RTP/M1 | Metrobus/Mexibus | Motorcycle taxi | Collective/micro | Bicycle | Light rail |
| Taxi app | Bus | Suburban | Personal transportation |  |  |  |
| Taxi | Motorcycle | Walk | Other |  |  |  |
| Subway | Trolleybus | Pedicab |  |  |  |  |

To update the O-D matrix, there are several methodologies in the literature. Most of them combine known information with current data observed such as the number of trips in some segments of the transit routes [15]. There are also approaches that project the trips to/from each zone based on the projected economic growth in those areas [55]. Nevertheless, given the pandemic situation we are experiencing today and that we have current available data about the increase or decrease in mobility for some modes of transport, transit stations and parking lots, we consider the 2017 O-D matrix as a reference matrix and update it to a scenario in 2020 using the daily mobility reports provided by Google [27] and the government of Mexico City [24]. According to [30], tables 3 to 6 represent the number of trips between these zones; for instance, the mean number of trips which origin is Coyoacán (id = 2) and destination is Iztapalapa (id = 6) during a week day is 228,272 (see Table 3) and the mean number of trips which origin is Tláhuac (id = 10) and destination is Cuauhtémoc (id = 14) is 21,881 (see Table 6) during a day of the weekend.

**Table 3:** Mean number of trips from zones 1-16 to zones 1-8 during a week day.

|  | 1 | 2 | 3 | 4 | 5 | 6 | 7 | 8 |
| --- | --- | --- | --- | --- | --- | --- | --- | --- |
| 1 | 477560 | 21108 | 7564 | 120676 | 6475 | 25026 | 1557 | 0 |
| 2 | 21111 | 886428 | 11010 | 48039 | 32230 | 228272 | 49305 | 17446 |
| 3 | 7336 | 10462 | 301805 | 9770 | 5397 | 10867 | 3491 | 722 |
| 4 | 120735 | 50796 | 9699 | 1677356 | 23427 | 52676 | 2602 | 590 |
| 5 | 6486 | 32393 | 5358 | 22936 | 361938 | 181660 | 1563 | 145 |
| 6 | 25244 | 221750 | 10308 | 56688 | 177845 | 2766471 | 9326 | 10395 |
| 7 | 1685 | 50941 | 4148 | 2576 | 1389 | 8008 | 260789 | 441 |
| 8 | 0 | 16022 | 722 | 590 | 145 | 10872 | 336 | 218175 |
| 9 | 24180 | 108129 | 102892 | 28836 | 18012 | 61990 | 80171 | 5577 |
| 10 | 1730 | 50969 | 1994 | 7283 | 8243 | 134119 | 2179 | 22801 |
| 11 | 11034 | 237367 | 10663 | 17958 | 14370 | 82757 | 53911 | 17203 |
| 12 | 2148 | 100785 | 2637 | 6062 | 7586 | 46856 | 7030 | 31284 |
| 13 | 17200 | 126058 | 14645 | 59944 | 59956 | 163732 | 29187 | 9714 |
| 14 | 88252 | 118292 | 20292 | 264298 | 84725 | 253810 | 22415 | 9495 |
| 15 | 99868 | 52246 | 43942 | 81013 | 26450 | 73629 | 11072 | 2972 |
| 16 | 15795 | 31971 | 3324 | 103651 | 56192 | 102397 | 3239 | 1199 |

**Table 4:**
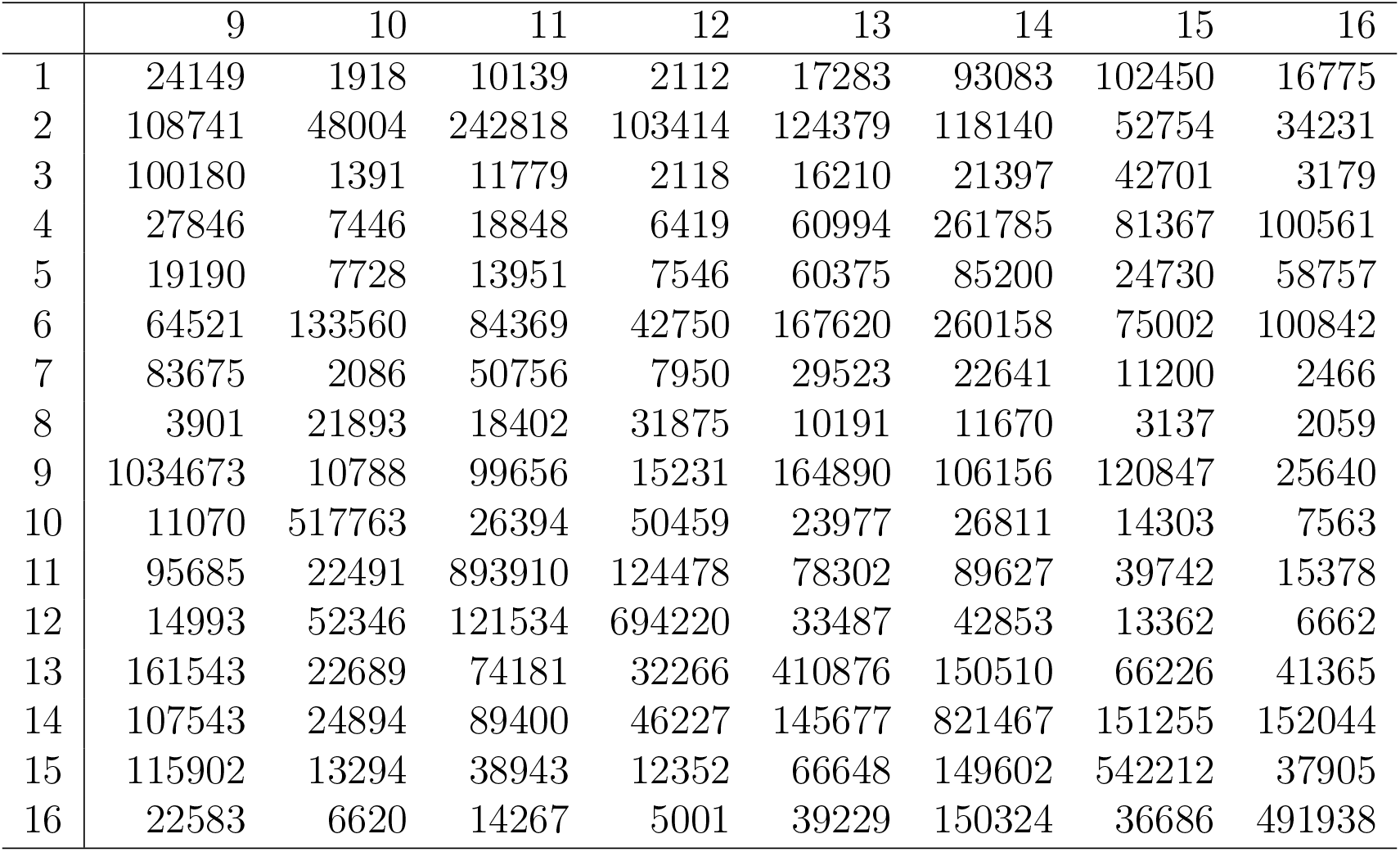
Mean number of trips from zones 1-16 to zones 9-16 during a week day.

**Table 5:**
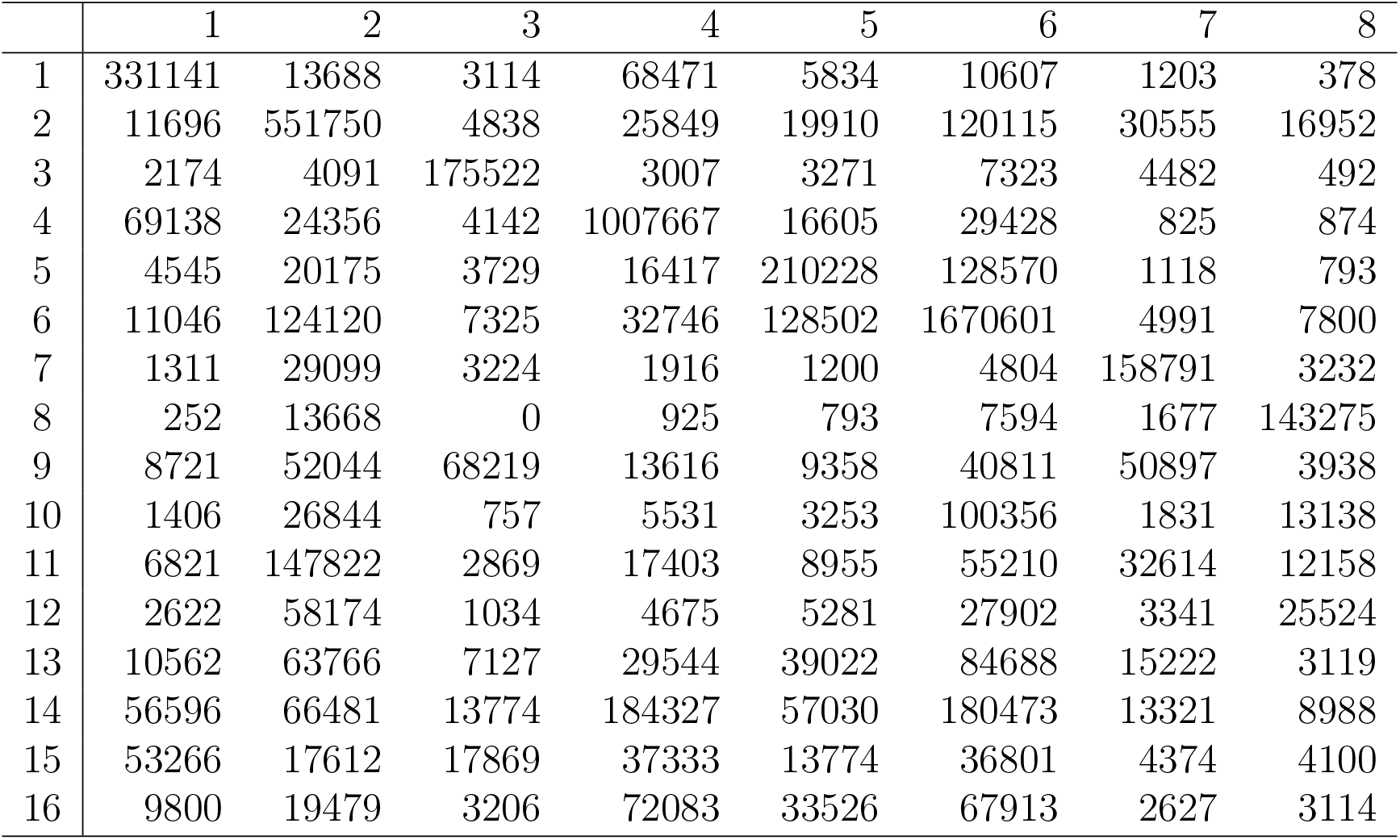
Mean number of trips from zones 1-16 to zones 1-8 during a day of the weekend.

|  | 1 | 2 | 3 | 4 | 5 | 6 | 7 | 8 |
| --- | --- | --- | --- | --- | --- | --- | --- | --- |
| 1 | 331141 | 13688 | 3114 | 68471 | 5834 | 10607 | 1203 | 378 |
| 2 | 11696 | 551750 | 4838 | 25849 | 19910 | 120115 | 30555 | 16952 |
| 3 | 2174 | 4091 | 175522 | 3007 | 3271 | 7323 | 4482 | 492 |
| 4 | 69138 | 24356 | 4142 | 1007667 | 16605 | 29428 | 825 | 874 |
| 5 | 4545 | 20175 | 3729 | 16417 | 210228 | 128570 | 1118 | 793 |
| 6 | 11046 | 124120 | 7325 | 32746 | 128502 | 1670601 | 4991 | 7800 |
| 7 | 1311 | 29099 | 3224 | 1916 | 1200 | 4804 | 158791 | 3232 |
| 8 | 252 | 13668 | 0 | 925 | 793 | 7594 | 1677 | 143275 |
| 9 | 8721 | 52044 | 68219 | 13616 | 9358 | 40811 | 50897 | 3938 |
| 10 | 1406 | 26844 | 757 | 5531 | 3253 | 100356 | 1831 | 13138 |
| 11 | 6821 | 147822 | 2869 | 17403 | 8955 | 55210 | 32614 | 12158 |
| 12 | 2622 | 58174 | 1034 | 4675 | 5281 | 27902 | 3341 | 25524 |
| 13 | 10562 | 63766 | 7127 | 29544 | 39022 | 84688 | 15222 | 3119 |
| 14 | 56596 | 66481 | 13774 | 184327 | 57030 | 180473 | 13321 | 8988 |
| 15 | 53266 | 17612 | 17869 | 37333 | 13774 | 36801 | 4374 | 4100 |
| 16 | 9800 | 19479 | 3206 | 72083 | 33526 | 67913 | 2627 | 3114 |

**Table 6:** Mean number of trips from zones 1-16 to zones 9-16 during a day of the weekend.

|  | 9 | 10 | 11 | 12 | 13 | 14 | 15 | 16 |
| --- | --- | --- | --- | --- | --- | --- | --- | --- |
| 1 | 9658 | 1501 | 5786 | 1542 | 9701 | 63331 | 53846 | 11181 |
| 2 | 52884 | 26855 | 150388 | 60418 | 64149 | 67953 | 19583 | 19995 |
| 3 | 65008 | 493 | 3714 | 856 | 8218 | 13863 | 18850 | 3176 |
| 4 | 11786 | 6370 | 16430 | 3851 | 28840 | 181498 | 37641 | 71621 |
| 5 | 8682 | 3140 | 9292 | 8080 | 40741 | 56400 | 13737 | 33730 |
| 6 | 43020 | 99372 | 52020 | 30083 | 94294 | 181288 | 40162 | 68541 |
| 7 | 51829 | 1013 | 37941 | 3235 | 14790 | 14315 | 4452 | 3892 |
| 8 | 2146 | 13264 | 10182 | 23451 | 2704 | 11629 | 3264 | 1256 |
| 9 | 579592 | 5619 | 54948 | 10135 | 87576 | 65512 | 60836 | 26414 |
| 10 | 6297 | 345171 | 17204 | 50337 | 14740 | 21881 | 6563 | 5956 |
| 11 | 55182 | 15078 | 546694 | 77342 | 41336 | 58766 | 21318 | 10691 |
| 12 | 9751 | 51898 | 76431 | 452836 | 17491 | 33051 | 8737 | 6028 |
| 13 | 85047 | 14753 | 38855 | 18884 | 232506 | 85671 | 28496 | 20327 |
| 14 | 60055 | 22350 | 60141 | 30310 | 84877 | 427114 | 92312 | 114306 |
| 15 | 59194 | 7156 | 19593 | 10801 | 27493 | 94657 | 339935 | 28238 |
| 16 | 22571 | 6200 | 10326 | 5581 | 20049 | 116777 | 26215 | 344904 |

To compute the new number of trips done using subway, RTP/M1, trolleybus, light rail or suburban, we used the corresponding rates given by the government of Mexico City; to compute the number of trips done using collective/micro or buses, we used the rates of transit stations given by Google and for personal transportation we used the workplaces rate. In the case of the trips done by bicycle, we used the rates for Ecobici and for metrobus/mexibus we used an average of both the rates of metrobus and mexibus in Mexico City. The rest of the transport modes remain the same as in 2017.

Figure 2 shows the variations in the mobility indices from February 27 to November 31, for each mode of transport that was modified.

**Figure 2:**
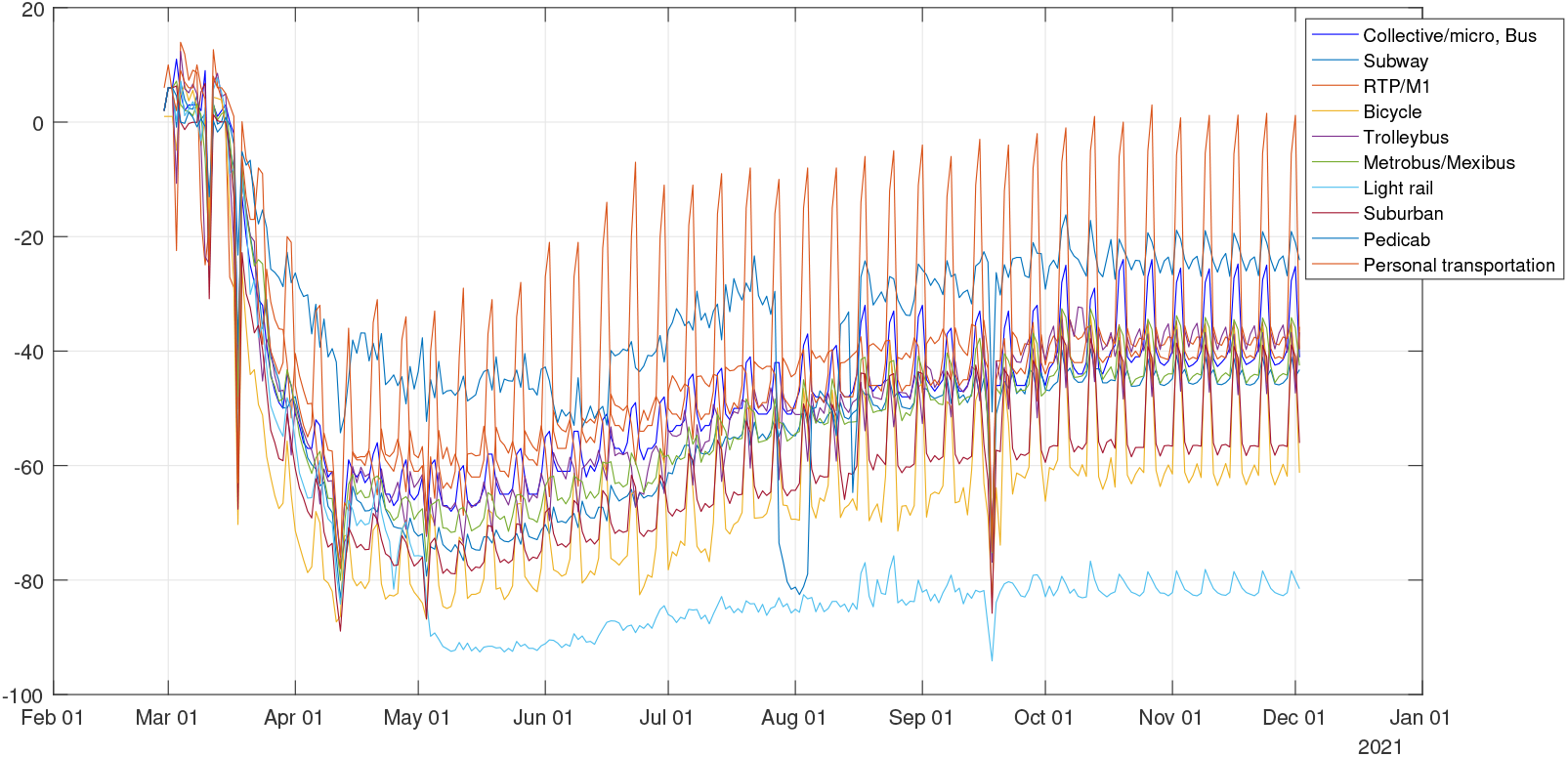
Boroughs of Mexico City.

In our mathematical model, the population of each borough is also considered. According to [31], those populations in 2020 are given in Table 7.

**Table 7:** Population in 2020 for each municipality of Mexico City.

| Id | Municipality | Population |
| --- | --- | --- |
| 1 | Azcapotzalco | 414 711 |
| 2 | Coyoacán | 628 063 |
| 3 | Cuajumalpa de Morelos | 199 224 |
| 4 | Gustavo A. Madero | 1 185 772 |
| 5 | Iztacalco | 384 326 |
| 6 | Iztapalapa | 1 815 786 |
| 7 | La Magdalena Contreras | 239 086 |
| 8 | Milpa Alta | 137 927 |
| 9 | Álvaro Obregón | 726 664 |
| 10 | Tláhuac | 305 076 |
| 11 | Tlalpan | 574 577 |
| 12 | Xochimilco | 407 885 |
| 13 | Benito Juárez | 385 439 |
| 14 | Cuauhtémoc | 531 831 |
| 15 | Miguel Hidalgo | 372 889 |
| 16 | Venustiano Carranza | 430 978 |

### 2.1 Clusters

In this section, we describe the clustering analysis implemented on Mexico City based on mobility data. During the pandemic, the Mexican government has scheduled four phases depending of level of contagion risk. These phases corresponding to the following periods: phase 1: from February 27th February to 22nd March; phase 2: from March 23rd to April 19th; phase 3: from April 20th to June 28th; phase 4: from June 29th to October 27th. The mobility network was analysed using the community detection module Louvain inside the *igraph* R package [23]. For more details about the *igraph* R package, see [22]. Thus, using the Louvain community detection algorithm, we identify that the Mexico City network has a modular structure, with 3 communities as shown in 3. These communities are the following: left top row shows the first phase of the pandemic. Community 1 is composed by boroughs 2, 3, 7, 8, 9, 11, 12 and 13, community 2 is composed by boroughs 1, 4, 15 and 16 and community 3 is composed by boroughs 5, 6 and 10. Right top row shows the second phase of the pandemic. Community 1 is composed by boroughs 3, 7, 9 and 13, community 2 is composed by boroughs 1, 4, 14, 15 and 16 and community 3 is composed by boroughs 2, 5, 6, 8, 10, 11 and 12. Left bottom row shows the third phase of the pandemic. Community 1 is composed by boroughs 3, 7, 9 and 13, community 2 is composed by boroughs 1, 4, 15 and 16 and community 3 is composed by boroughs 2, 5, 6, 8, 10, 11 and 12. Right bottom row shows the fourth phase of the pandemic. Community 1 is composed by boroughs 2, 3, 7, 8, 9, 11, 12 and 13, community 2 is composed by boroughs 1, 4, 15 and 16 and community 3 is composed by boroughs 5, 6 and 10.

We also have done an clustering analysis for a bigger region, the Metropolitan Area of Valley of Mexico (MAVM), the most affected region of diagnosed cases of coronavirus in Mexico country in order to explore which boroughs of Mexico City share the same community with municipalities of Mexico State. The MAVM consists of all the 16 boroughs of Mexico City, 59 municipalities of the State of Mexico and 1 municipality of the state of Hidalgo. Here, we have taken into account the eighteen most representative municipalities of Mexico State. Thus, we considered a sub-area of MAVM consisting of a total of 34 zones. The municipalities considered of Mexico State have the following identification Id: Coacalco de Berriozabal, Cuautitlan, Chalco, Chicoloapan, Chimalhuacan, Ecatepec de Morelos, Huixquilucan, Ixtapaluca, Naucalpan de Juarez, Nezahualcoyotl, Nicolas Romero, La Paz, Tecamac, Texcoco, Tlalnepantla de Baz, Tultitlan, Cuautitlan Izcalli, Balle de Chalco Solidaridad, 17, 18, 19, 20, 21, 22, 23, 24, 25, 26, 27, 28, 29, 30, 31, 32, 33, and 34, respectively. Figure 4 shows that the sub-area of MAVM is divided in four communities based on mobility data. These communities remain the same during the four phases. The communities are the following: community 1 is composed by boroughs 2, 3, 5, 6, 7, 8, 9, 10, 11, 12, 13 and 28, community 2 is composed by boroughs 1, 15, 17, 18, 23, 25, 27, 31, 32 and 33, community 3 is composed by boroughs 4, 14, 16, 20, 21, 22, 26, 29 and 30 and finally community 4 is composed by boroughs 19, 29 and 34. Figure 4 shows that the boroughs 1, 4, 14, 15, and 16 of Mexico City belong to the communities 2 and 3 which are not part of the main community of the Mexico City, i.e., boroughs 1 and 15 belong to the community 2 and boroughs 4, 14 and 16 belongs to the community 3. Therefore, based on the origin-destination matrices, we can see that the boroughs 1, 4, 14, 15 and 16 of Mexico City are more communicated with the Mexico State area. This could be explained in part geographically since the boroughs 1, 4, 14, 15 and 16 are in the north of Mexico City and particularly the boroughs 1, 4, 15 and 16 are on the boundary between Mexico City and Mexico State, secondly, the explanation has to do that people from Mexico State travel on daily basis to these boroughs 4, 14, 15 and 16 to work there since these boroughs concentrate business centres and corporation headquarters. From figure 4, we also can observe that the borough 28 (La Paz) from Mexico State is more connected with the Mexico City area, this may be due to the fact that it borders to the west and south-west with one of the most populated municipalities in Mexico City, Iztapalapa, and that it connects directly with three subway lines.

**Figure 3:**
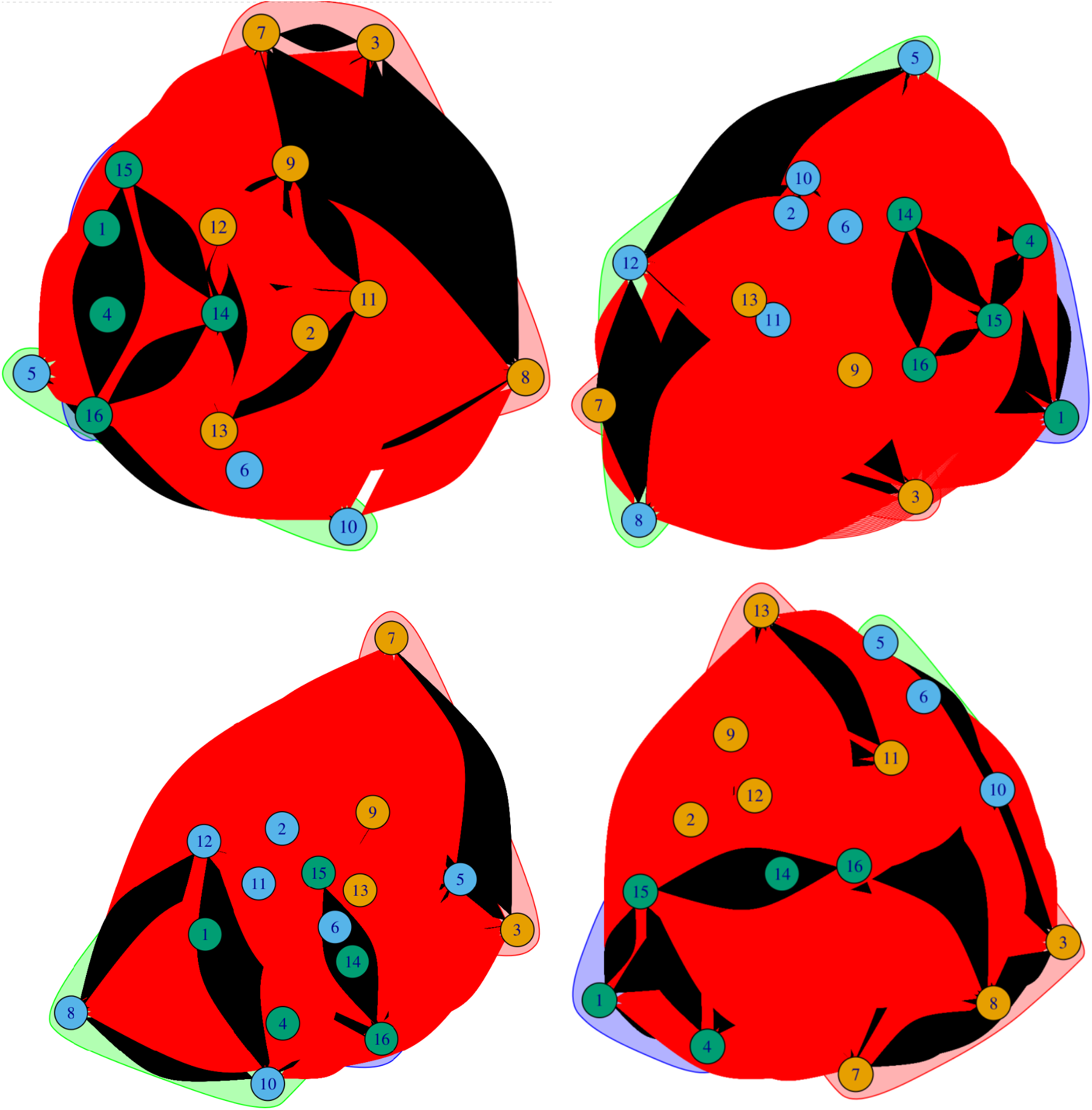
District clusters of Mexico City. Top row from left to right: the District clusters for the first period of the pandemic, and the Districts clusters of the second period of the pandemic. Bottom row from left to right: the District clusters of the third and the fourth periods of the pandemic

**Figure 4:**
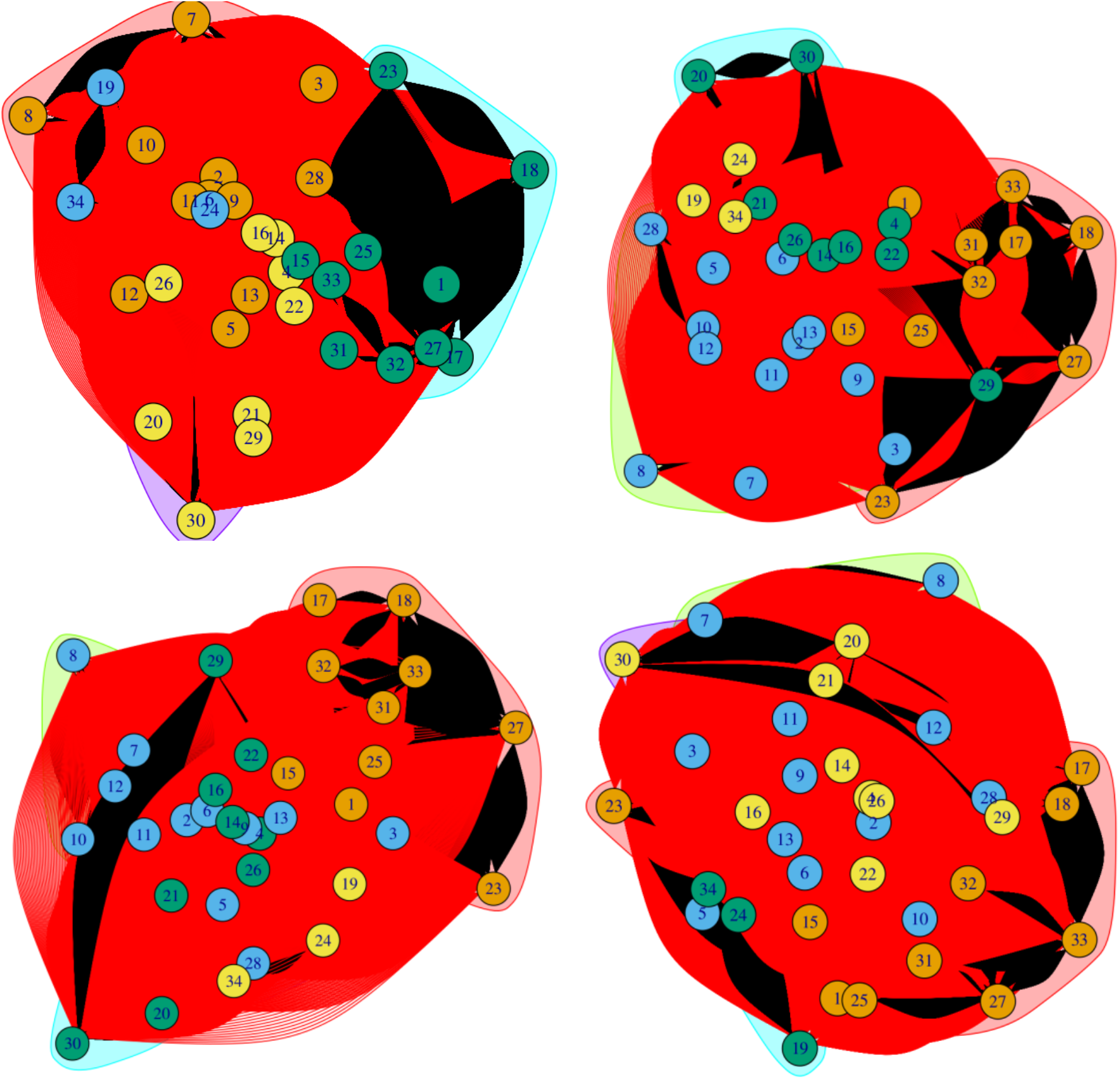
District clusters of Valle de Mexico. Top row from left to right: the District clusters of the first period of the pandemic, and the Districts clusters of the second period of the pandemic. Bottom row from left to right: the District clusters of the third and the fourth periods of the pandemic

## 3 Mathematical model

As we mention in the Introduction, the transmission model incorporates information on human movement within the following Susceptible, Exposed, Infected, Recovered (SEIR) metapopulation structure [35]:

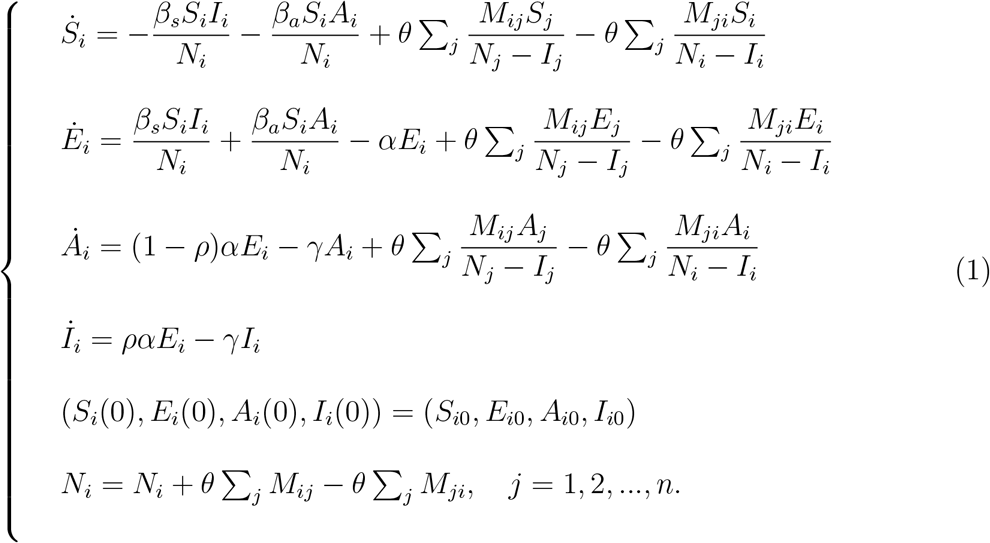

In above model the symbol ·means the derivative in time. Additionally, *S*_*i*_, *E*_*i*_, 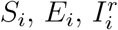 and 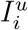 are the susceptible, exposed, documented infected, undocumented infected being *N*_*i*_ the total population in city *i* at time *t*. Also, it is assumed that individuals in the 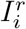 group do not move between cities, though these individuals can move between cities during the latency period.

A complete description of the parameters involved in the model (1), the respective range of values proposed in [35] and their measurement units can be found on Table 8.

**Table 8:** ModelParameters.

|  |  |  |  |
| --- | --- | --- | --- |
| $\beta_s$ | Transmission rate due to documented infected individuals. | $[0., 1.]$ | None |
| $\beta_a$ | Transmission rate due to undocumented individuals. | $[0., 1.]$ | None |
| $\frac{1}{\theta}$ | Multiplicative factor (*reflect under-reporting of human movement*). | $[0., 1.]$ | *trips/person* |
| $\frac{1}{\alpha}$ | Average latency period. | $[2, 10]$ | *days* |
| $\frac{1}{\gamma}$ | Average duration of infection. | $[2, 20]$ | *days* |
| $\rho$ | Fraction undocumented/documented people. | $[0., .4]$ | None |

**Table 9:** Parameter estimation of *β*_*s*_, *β*_*a*_, *ρ, α, γ, θ* and initial conditions of the model (1).

|  | mean | std | min | 2.5% | 50% | 97.5% | max |
| --- | --- | --- | --- | --- | --- | --- | --- |
| $\beta_s$ | 0.1874 | 0.0970 | 0.0252 | 0.0561 | 0.1674 | 0.4237 | 0.5859 |
| $\beta_a$ | 0.2653 | 0.0714 | 0.0837 | 0.1413 | 0.2607 | 0.4027 | 0.5472 |
| $\rho$ | 0.0275 | 0.0073 | 0.0108 | 0.0160 | 0.0266 | 0.0441 | 0.0563 |
| $\alpha$ | 0.1675 | 0.0436 | 0.1023 | 0.1120 | 0.1575 | 0.2783 | 0.3721 |
| $\gamma$ | 0.2057 | 0.0568 | 0.0761 | 0.1072 | 0.2015 | 0.3175 | 0.3923 |
| $\theta$ | 0.2607 | 0.0644 | 0.0995 | 0.1487 | 0.2543 | 0.3972 | 0.4845 |
| $E_{10}$ | 184.3217 | 146.0019 | 3.3820 | 20.2310 | 143.0350 | 555.9260 | 882.5690 |
| $A_{10}$ | 535.6228 | 262.4267 | 18.3910 | 58.9131 | 544.1240 | 945.0320 | 996.0400 |
| $I_{10}$ | 3.3211 | 4.1348 | 0.0710 | 0.2166 | 2.0098 | 14.4367 | 54.8012 |
| $E_{20}$ | 205.7978 | 169.3601 | 5.8993 | 19.8599 | 160.9300 | 643.1670 | 889.7030 |
| $A_{20}$ | 209.4122 | 170.9916 | 2.4569 | 16.9377 | 159.3900 | 654.5110 | 900.7740 |
| $I_{20}$ | 2.9190 | 3.1590 | 0.0865 | 0.3008 | 1.9768 | 11.1929 | 31.2103 |
| $E_{30}$ | 145.8133 | 135.1257 | 2.7717 | 12.4263 | 98.9493 | 520.7570 | 777.5560 |
| $A_{30}$ | 402.3670 | 245.8086 | 6.1237 | 40.3436 | 375.2480 | 894.0940 | 990.3010 |
| $I_{30}$ | 4.3542 | 5.5809 | 0.0540 | 0.2546 | 2.5353 | 19.0474 | 55.5592 |
| $E_{40}$ | 702.9435 | 273.8195 | 5.7849 | 86.0434 | 794.9580 | 991.9220 | 999.3340 |
| $A_{40}$ | 432.1857 | 273.8040 | 10.5007 | 42.6838 | 403.6190 | 913.8580 | 990.9040 |
| $I_{40}$ | 35.8966 | 30.9604 | 0.0317 | 0.5399 | 27.3064 | 95.5256 | 99.6371 |
| $E_{50}$ | 537.4395 | 276.9246 | 7.5696 | 55.8129 | 551.4940 | 961.3700 | 999.5670 |
| $A_{50}$ | 464.0044 | 282.1865 | 0.7563 | 34.5782 | 432.4700 | 956.6170 | 990.0100 |
| $I_{50}$ | 5.4617 | 6.9190 | 0.0645 | 0.2225 | 2.8948 | 25.1745 | 51.6870 |
| $E_{60}$ | 449.9065 | 295.9580 | 0.3066 | 23.2522 | 415.6940 | 956.0590 | 996.7500 |
| $A_{60}$ | 432.2875 | 289.1433 | 2.7484 | 24.1510 | 395.6120 | 962.4810 | 995.5330 |
| $I_{60}$ | 26.2795 | 27.2295 | 0.0364 | 0.3178 | 14.9401 | 90.2088 | 98.8098 |
| $E_{70}$ | 83.9107 | 79.6407 | 2.3274 | 8.9936 | 60.3275 | 296.4430 | 643.2260 |
| $A_{70}$ | 476.9134 | 267.1927 | 1.8749 | 57.2647 | 468.6710 | 941.6410 | 989.7840 |
| $I_{70}$ | 2.8198 | 3.5169 | 0.0306 | 0.1553 | 1.6862 | 13.0140 | 47.4353 |
| $E_{80}$ | 190.3153 | 165.8991 | 2.2265 | 16.1677 | 138.7190 | 632.7900 | 907.4100 |
| $A_{80}$ | 513.3394 | 300.4417 | 1.3576 | 27.0607 | 523.6620 | 970.2510 | 997.5210 |
| $I_{80}$ | 3.0580 | 5.3752 | 0.0239 | 0.1848 | 1.4966 | 13.3490 | 87.2188 |
| $E_{90}$ | 180.8683 | 163.3549 | 2.7683 | 11.5561 | 130.9020 | 613.8010 | 903.1110 |
| $A_{90}$ | 689.9932 | 264.9430 | 0.9175 | 112.3040 | 773.1550 | 989.1490 | 998.4900 |
| $I_{90}$ | 16.5314 | 19.5683 | 0.0135 | 0.2542 | 8.4786 | 70.1495 | 98.5526 |
| $E_{100}$ | 298.3463 | 237.1295 | 2.6486 | 17.2592 | 232.3220 | 837.6510 | 955.7100 |
| $A_{100}$ | 623.9403 | 319.4492 | 1.7870 | 24.2471 | 713.8970 | 994.4600 | 999.7380 |
| $I_{100}$ | 36.2087 | 34.8954 | 0.0002 | 0.1429 | 21.8228 | 98.7000 | 99.9149 |
| $E_{110}$ | 243.8098 | 214.7284 | 2.0004 | 11.5777 | 175.9570 | 784.3620 | 968.7030 |
| $A_{110}$ | 198.8057 | 180.8828 | 2.1306 | 12.4892 | 137.6340 | 679.0910 | 910.9480 |
| $I_{110}$ | 5.1452 | 7.4224 | 0.0287 | 0.1621 | 2.4138 | 27.1594 | 61.6416 |
| $E_{120}$ | 97.3070 | 100.6553 | 1.5153 | 7.4905 | 63.6840 | 383.0570 | 734.7550 |
| $A_{120}$ | 156.2853 | 174.5743 | 0.3561 | 4.7232 | 85.3196 | 642.8160 | 972.1780 |
| $I_{120}$ | 6.7454 | 9.5071 | 0.0206 | 0.1810 | 3.2779 | 35.0406 | 79.3457 |
| $E_{130}$ | 103.1659 | 114.1921 | 2.0040 | 5.4339 | 60.5820 | 436.4240 | 803.5820 |
| $A_{130}$ | 149.8156 | 146.1935 | 1.6622 | 8.8127 | 97.8790 | 582.2150 | 775.5250 |
| $I_{130}$ | 17.5729 | 23.3707 | 0.0052 | 0.1023 | 6.2487 | 84.1743 | 99.4204 |
| $E_{140}$ | 218.2782 | 210.3152 | 1.4959 | 7.4696 | 141.5680 | 773.9480 | 955.9370 |
| $A_{140}$ | 99.6862 | 121.2945 | 1.2440 | 4.4774 | 51.7817 | 478.1060 | 705.4500 |
| $I_{140}$ | 1.7250 | 2.3882 | 0.0168 | 0.1084 | 0.9497 | 8.2495 | 30.9248 |
| $E_{150}$ | 153.4127 | 149.2736 | 1.4227 | 9.8697 | 101.7880 | 544.8440 | 826.6820 |
| $A_{150}$ | 557.2493 | 333.0496 | 0.5477 | 14.0250 | 599.8180 | 993.3000 | 999.0760 |
| $I_{150}$ | 36.5668 | 34.1058 | 0.0039 | 0.1421 | 25.0996 | 99.2558 | 99.9876 |
| $E_{160}$ | 559.8946 | 344.4055 | 0.4518 | 8.6621 | 617.1160 | 997.2990 | 999.8600 |
| $A_{160}$ | 457.4253 | 338.2471 | 0.1368 | 5.4778 | 428.0480 | 985.6480 | 999.3850 |
| $I_{160}$ | 36.5718 | 32.8368 | 0.0210 | 0.3451 | 26.4229 | 97.0763 | 99.6811 |

**Table 10:**
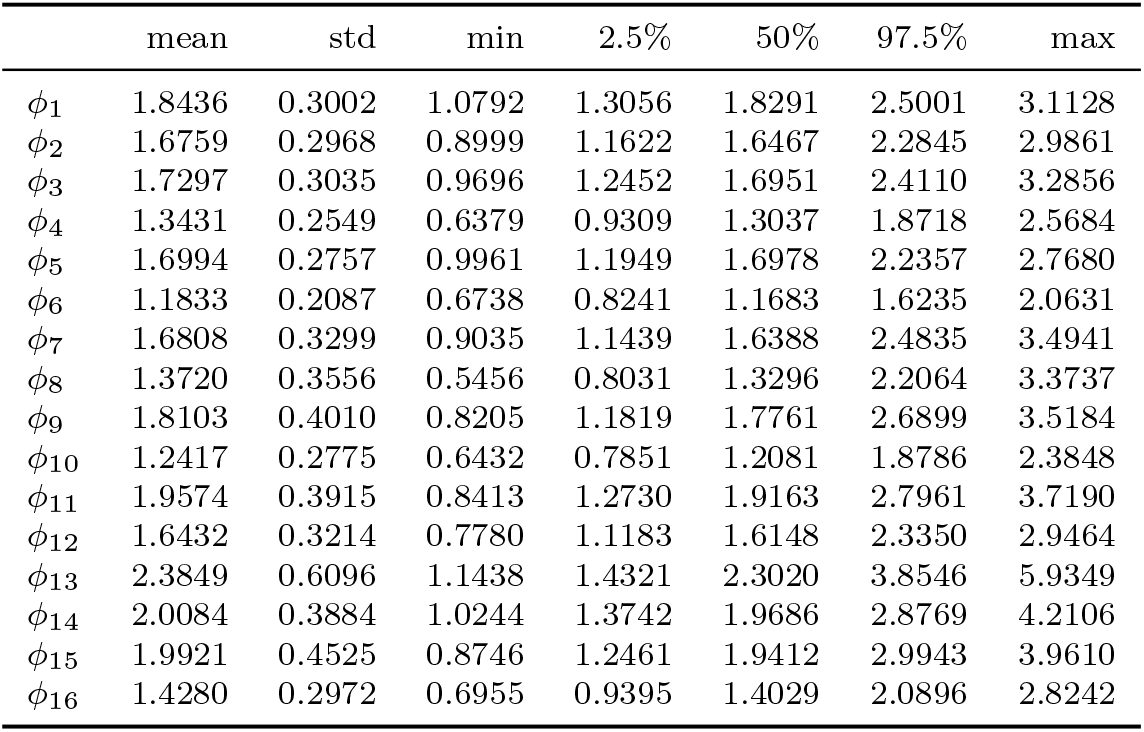
Over-dispersion parameters’ estimation of the Negative Binomial Distribution (7).

## 4 The basic reproduction number estimation

The basic reproduction number, commonly denoted by ℛ_0_ is the average number of secondary infections generated by a single infective during the curse of the infection in a whole susceptible population. We calculate the reproductive number R_*e*_ in Mexico City using the inferred parameters. Define **X** = (*E, A, I*) and using the next generation operator method [26] on the system (1), the Jacobian matrices ℱ and 𝒱 of system (1) are given by

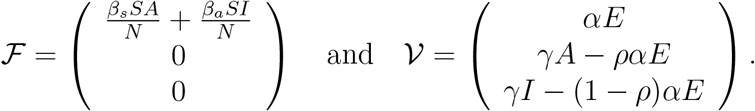

The disease free equilibrium (DFE) of system (1) is **X**_0_ = (0, 0, 0, *N*, 0)^*T*^, then we have

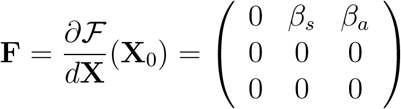

and

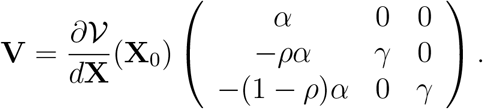

Therefore, the next-generation matrix is **K** = **FV**^−1^, from where *R*_*e*_ is computed as the leading eigenvalue of matrix **K**. i.e,

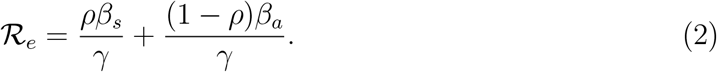

Note that at the beginning of the epidemic (e.g before February 27, 2020), ℛ_*e*_ is equivalent to ℛ_0_. As the epidemic unfolds, a declination of susceptible population reduces the effective reproductive number. Here, we used the same formula above for periods after February 27, 2020.

Table 8 shows the range of values for the parameters involved in the expression (2) obtained using the Stan package [13]. With those values we obtain a %95 credible interval for ℛ_*e*_ ∈ (1.2713, 1.3054).

## 5 Parameter estimation

For parameter estimation we use the daily reported data-set [25]. For this end, we use Bayesian inference to solve the inverse problem associated to the system of Ordinary Differential Equations (ODEs) given on (1) similarly to [47]. Some references using this method of parameter estimation can be found in [1, 3, 6, 7, 8, 9, 11, 14, 18, 28, 37, 56].

Let us denote the vector of state variables in the zone *i* as **x** = (*S*_*i*_, *E*_*i*_, *A*_*i*_, *I*_*i*_) ∈ (*L*^2^[0, *T*])^*n*^, where *n* = 4 denotes the number of state variables and the vector of parameters in the zone *i* as 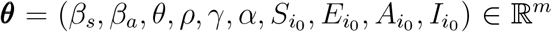, where *m* = 10 denotes the dimension number of parameters to estimate. Thus, we can write the model (1) as the following Cauchy problem

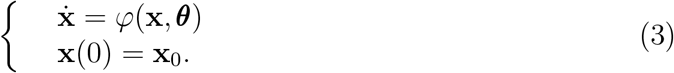

Problem (3), defines a mapping Φ(***θ***) = **x** from parameters ***θ*** to state variables **x**, where 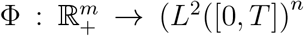, where ℝ_+_ denotes the non-negative real numbers. We assume that Φ has a Fréchet derivative. Therefore, the inverse problem is formulated as a standard optimization problem

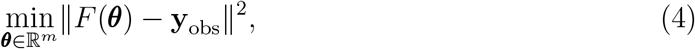

such that *x* = Φ(***θ***) holds, with **y**_obs_ is the data which has error measurements of size *η*. Problem (3) may be solved using numerical tools to deal with a non-linear least-squares problem [2, 12, 48, 49, 54]. In this work, we implement Bayesian inference to solve the inverse problem given on (4). From the Bayesian perspective, all state variables **x** and parameters ***θ*** are considered as random variables and the data **y**_obs_ is fixed. For the random variables **x** and ***θ***, the joint probability distribution density of the data **x** and the parameters ***θ***, denoted by *π*(***θ*, x**), is given by *π*(***θ*, x**) = *π*(**x**|***θ***)*π*(***θ***), where *π*(**x**|***θ***)*π*(***θ***) is the conditional probability distribution, also called the likelihood function, and *π*(**x**|***θ***) is the prior distribution which involves the prior information of parameters ***θ***. Given **x** = **y**_obs_, the conditional probability distribution *π*(***θ***|**y**_obs_), called the posterior distribution of ***θ*** is given by the Bayes’ theorem:

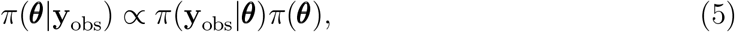

If an additive noise is assumed

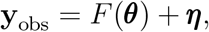

where ***η*** is the noise due to discretization, the model error and the measurement error. If the noise probability distribution *π*_*H*_ (***η***) is known, ***θ*** and ***η*** are independent, then

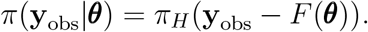

All the available information regarding the unknown parameter ***θ*** is codified into the a prior distribution *π*(***θ***), it specifies our belief in a parameter before observing the data. All the available information regarding the way of how was obtained the measured data is codified into the likelihood distribution *π*(**y**_obs_|***θ***). This likelihood can be seen as an objective or cost function, as it punishes deviations of the model from the data. To solve the associated inverse problem (5), one may use the maximum a posterior (MAP)

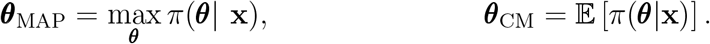

We used the data set in the zone *i* as 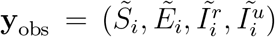, which correspond to the susceptible, exposed, documented infected and undocumented infected in the z *i*, respec-tively. A Poisson distribution with respect to the time is typically used to account for the discrete nature of these counts. However, the variance of each component of the data set **y**_obs_ is larger than its mean, which indicates that there is over-dispersion of the data. Thus, a more appropriate likelihood distribution is to use the Negative Binomial (NB), since it has an additional parameter that allows the variance to exceed the mean [3, 11, 42]. In fact the NB is a mixture of Poisson and Gamma distributions, where the rate parameter of the Poisson distribution itself follows a Gamma distribution [21, 42]. We mention there exists different mathematical expressions for the NB depending on the author or source, they are equivalent. Because of this multiple representation of the NB in the literature, one must assure to use the NB distribution accordingly to the source. Here, we have used the following expression for the NB distribution

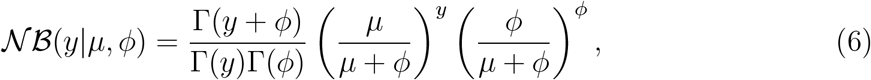

where *µ* is the mean of the random variable *y* ∼ 𝒩 ℬ (*y*|*µ, ϕ*) and *ϕ* is the over-dispersion parameter, i.e.,

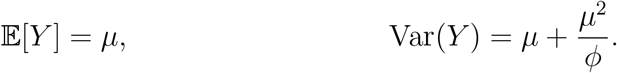

We recall that the Poisson distribution has mean and variance equal to *µ*, so *µ*^2^*/ϕ >* 0 is the additional variance of the NB with respect to the Poisson distribution. The inverse of the parameter *ϕ*, controls the over-dispersion, thus, it is important selecting its support adequately for parameter estimation. Also, there exists alternative forms of the NB distribution. In fact, we have used the first option *neg bin* of the NB distribution of Stan [13]. We acknowledge that some scientists have had success with the second alternative representation of the NB distribution [28]. We assume independent NB distributed noise ***η***, i.e., all dependency in the data is codified into the contact tracing model. In other words, the positive definite noise covariance matrix ***η*** is assumed to be diagonal. Therefore, using the Bayes formula, the likelihood is

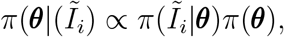

where *i* denotes the borough index. As mentioned above, we approximate the likelihood probability distribution corresponding to diagnosed cases with a NB distribution

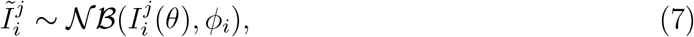

where the index *j* denotes the number of days, *i* the number of the boroughs, and *ϕ*_*i*_ are the parameters corresponding to the over-dispersion parameter of the NB distribution (6) respect to each borough. For independent observations, the likelihood distribution *π*(**y** | ***θ***), is given by the product of the individual probability densities of the observations

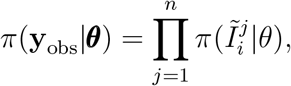

where the mean *µ* of the NB distribution 𝒩 ℬ (*I*_*i*_(*θ*), *ϕ*_*i*_), is given by the solution *I*_*i*_(*t*) of the model (1) at time *t* = *t*_*j*_. For the prior distribution, we select the `LogNormal` distribution for *β*_*s*_ and *β*_*a*_ parameters, `Gamma` distributions for *α* and *γ* parameters and

Uniform distributions for the rest of parameters to estimate: *ρ, θ* and initial conditions 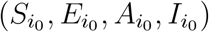. The hyperparameters and their support corresponding to all the distributions of the parameters to estimate are given on table’s range 8.

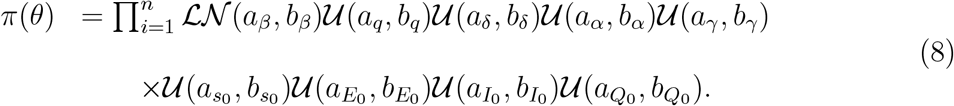

The posterior distribution *π*(*θ* |*y*_obs_) given by (5) does not have an analytical closed form since the likelihood function, which depends on the solution of the non-linear model given on (1), does not have an explicit solution. Then, we explore the posterior distribution using two methods: the Stan Statistics package [13] within its version the Automatic Differentiation Variational Inference (ADVI) method, and secondly, the general purpose Markov Chain Monte Carlo Metropolis-Hasting (MCMC-MH) algorithm `t-walk` [19]. Both algorithms generate samples form the posterior distribution *π*(***θ***|**y**_obs_) that can be used to estimate marginal posterior densities, mean, credible intervals, percentiles, variances, and others. We refer to [29] for a more complex MCMC-MH algorithms.

Figure 5 shows the credible intervals of parameters of model (1) within 95% Highest-Posterior Density (HPD) using the Stan Package [13]. Figures 6-8 show the fit of confirmed COVID-19 cases of all the boroughs of Mexico City using the Stan [13]. Figure 10 shows Credible intervals of parameters of model (1) within 95% Highest-Posterior Density (HPD) using the ADVI-Meanfield method of Stan package [13]. Figure 11 shows Credible intervals of parameters of model (1) within 95% Highest-Posterior Density (HPD) using the t-walk Package [19]. We point out that results obtained with the t-walk package are preliminary since we performed only 60,000 iterations with 30,000 of them as burn-in. We performed this limited quantity of iterations since the computational time consumption is significantly large for each 1,000 of iterations. We will perform more iterations in the near future. Using both packages, we did a fit for the first 245 days of the pandemic in Mexico City, starting the 27th of February, and we have performed predictions from 245 − 275 day, corresponding to October 28-th to November 28-th. We set up an minimum borough fraction equal to 0.6 to limit the borough to fall below their population size. Some future work will correspond to analyse the identifiability of the parameters of model (1), as suggested in [18, 39, 50], specifically the *ρ* parameter since this parameter is multiplied by the period of incubation of the disease, *α*, thus, estimating both parameters simultaneously may lead to non-identifiability difficulty.

**Figure 5:**
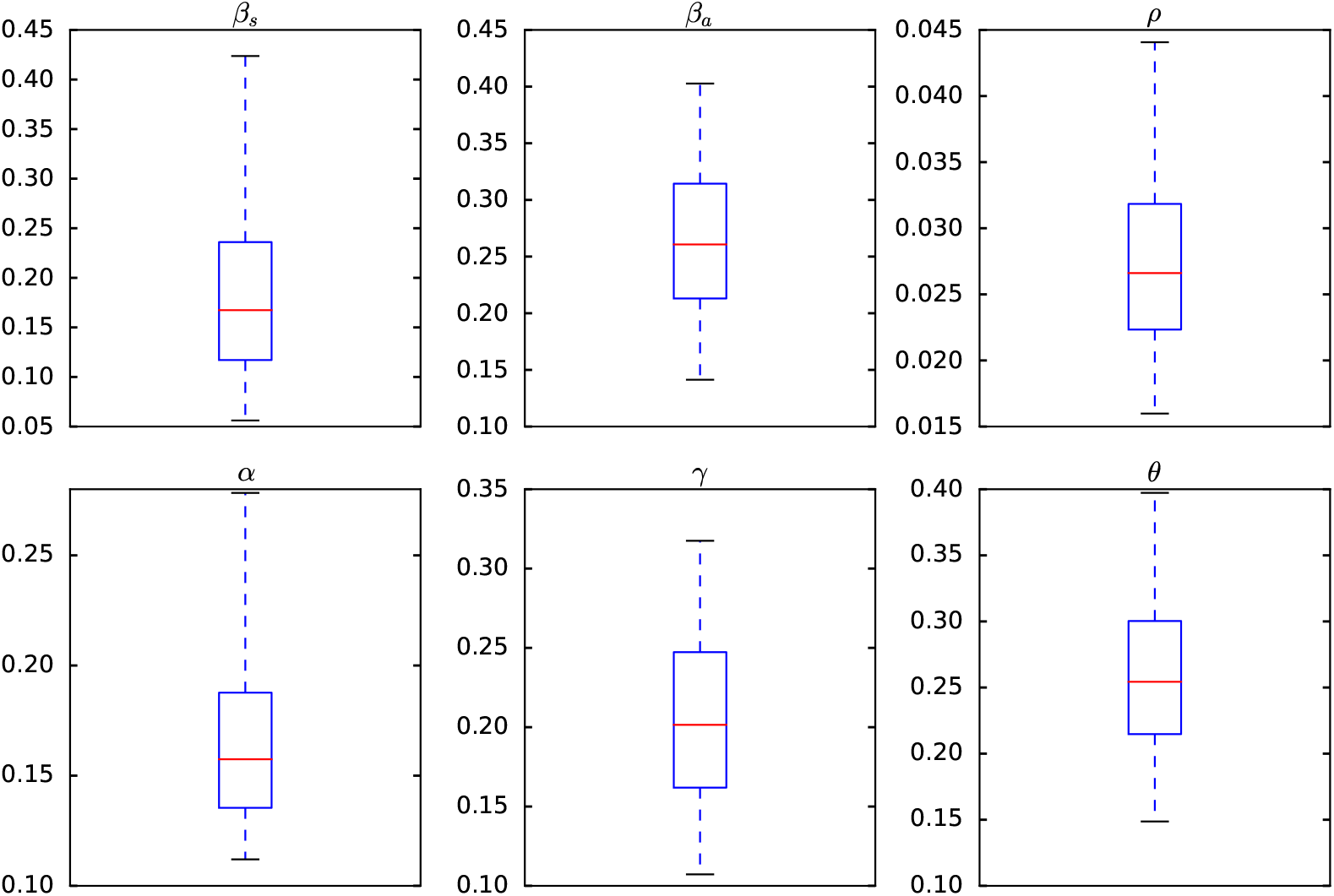
Credible intervals of parameters of model (1) within 95% Highest-Posterior Density (HPD) using the Stan Package [13]

**Figure 6:**
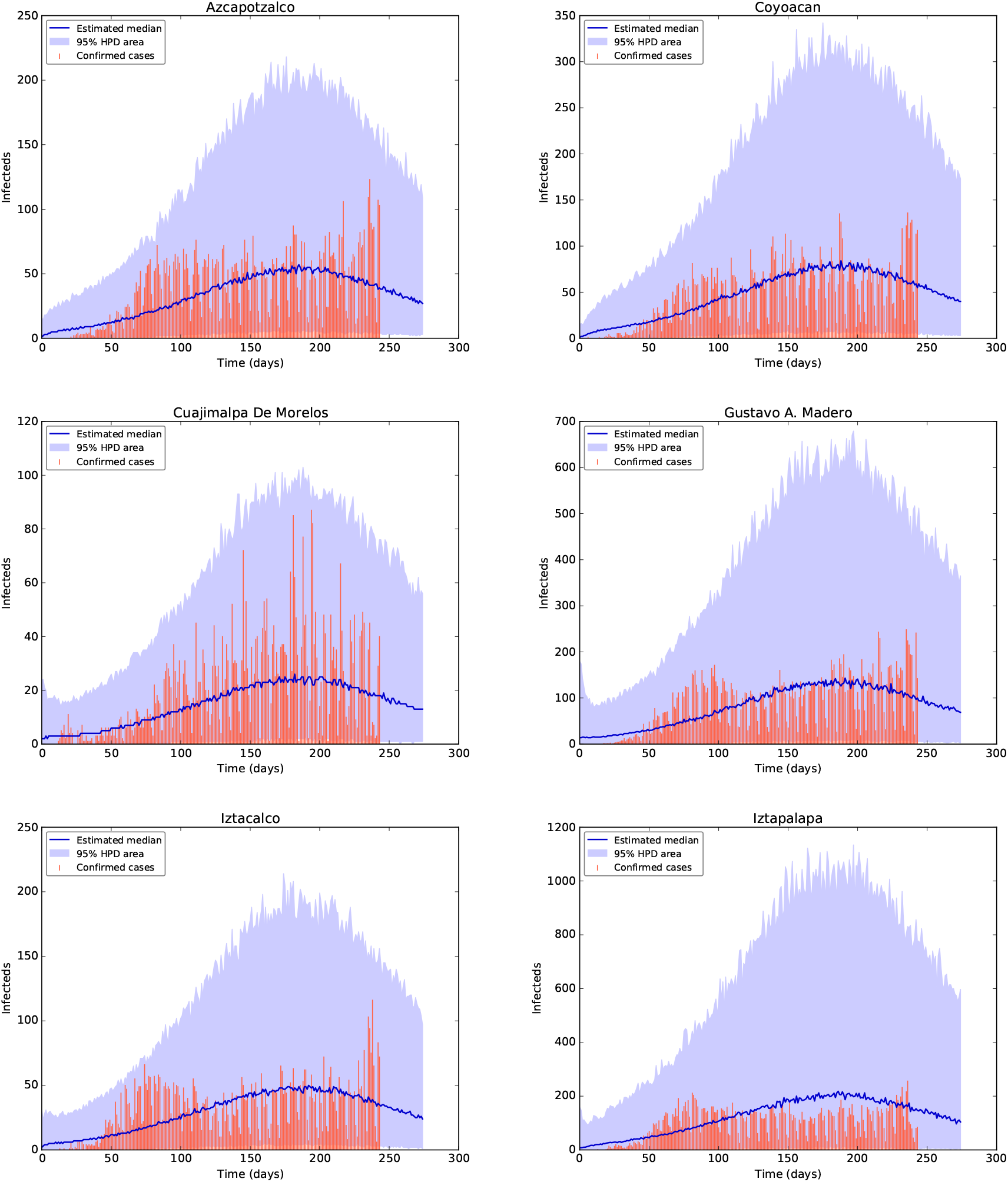
Fit of confirmed COVID-19 cases using the Stan package [13]. Top row from left to right: the fit for the confirmed cases of the Districts Azcapotzalco and Coyoacan. The tomatoe colour bars reprent the confirmed cases, the blue and purple solid lines reprent the median and the mode, respectively, and the shaded area represent the %95 probability bands for the expected value for the state variable of Documented Infecteds. Middle row from left to right: the fit for the diagnosed cases of the Districts Cuajimalpa de Morelos and Gustavo A. Madero. Bottom row from left to right: the fit for the diagnosed cases of the Districts Iztacalco and Iztapalapa.

**Figure 7:**
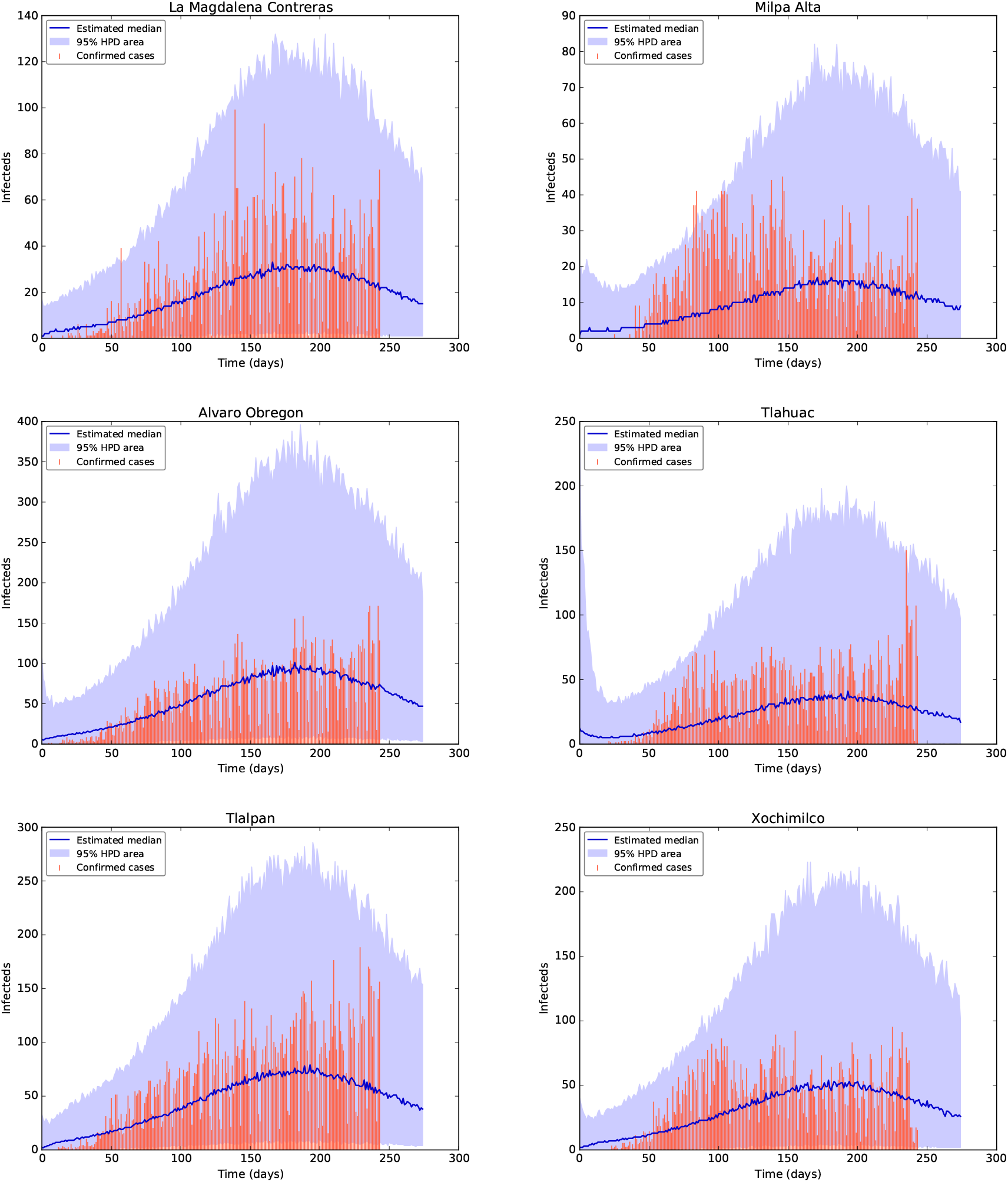
Fit of confirmed COVID-19 cases using the Stan package [13]. Top row from left to right: the fit for the confirmed COVID-19 cases of the Districts La Magdalena Contreras and Milpa Alta. The tomatoe colour bars reprent the confirmed COVID-19 cases, the blue solid line reprent the median and the shaded area represent the %95 probability bands for the expected value for the state variable of Documented Infecteds. Middle row from left to right: the fit for the diagnosed cases of the Districts Alvaro Obregon and Tlahuac. Bottom row from left to right: the fit for the diagnosed cases of the Districts Tlalpan and Xochimilco.

**Figure 8:**
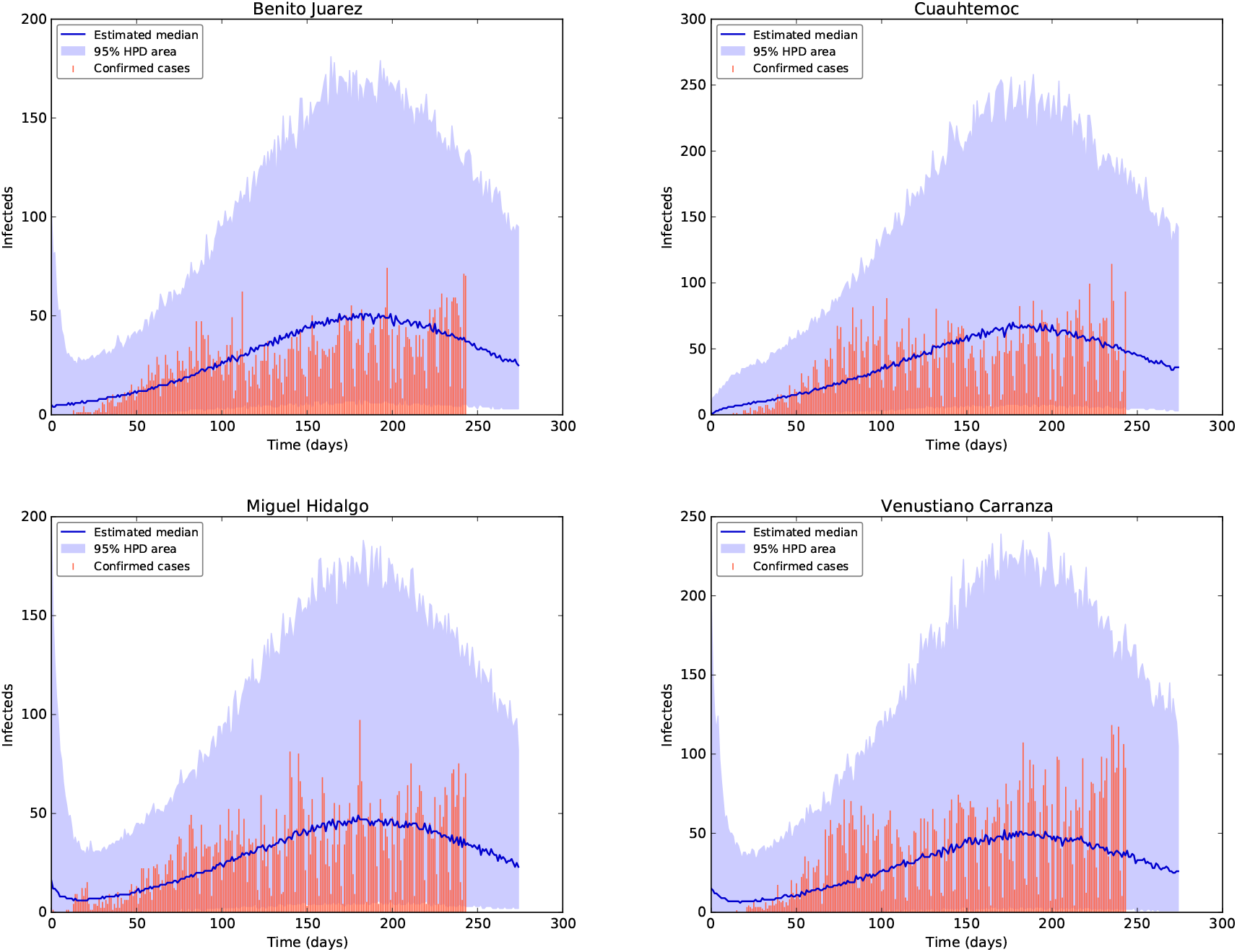
Fit of confirmed COVID-19 cases using the Stan package [13]. Top row from left to right: the fit for the confirmed COVID-19 cases of the Districts Benito Juarez and Cuauhtemoc. The tomatoe colour bars reprent the confirmed COVID-19 cases, the blue solid line reprent the median and the shaded area represent the %95 probability bands for the expected value for the state variable of Documented Infecteds. Bottom row from left to right: the fit for the diagnosed cases of the Districts Miguel Hidalgo and Venustiano Carranza.

**Figure 9:**
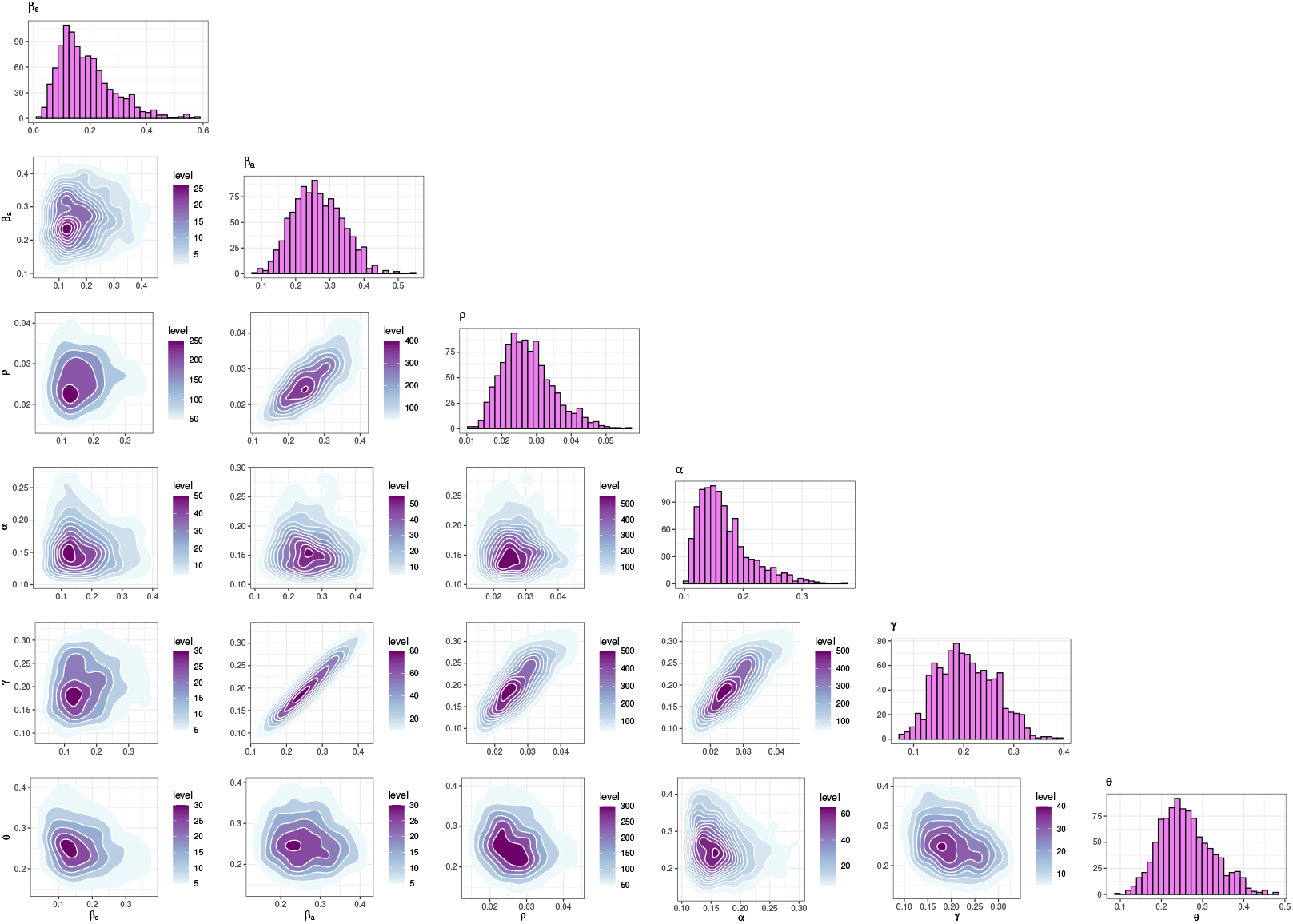
Joint probability density distributions of the estimated parameters within 95% (HPD). The blue lines represent the medians.

**Figure 10:**
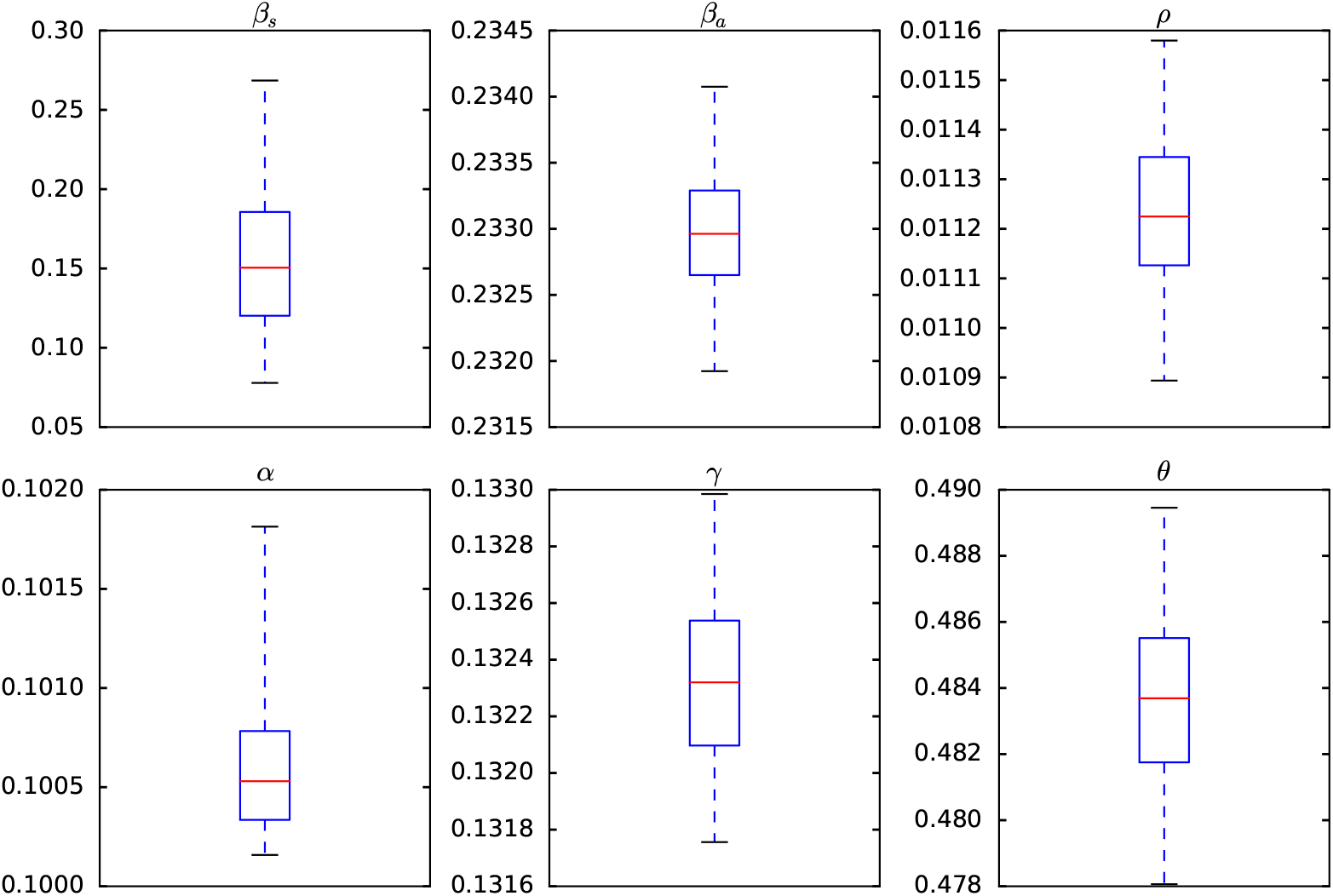
Credible intervals of parameters of model (1) within 95% Highest-Posterior Density (HPD) ADVI-Meanfield method of Stan package [13]

**Figure 11:**
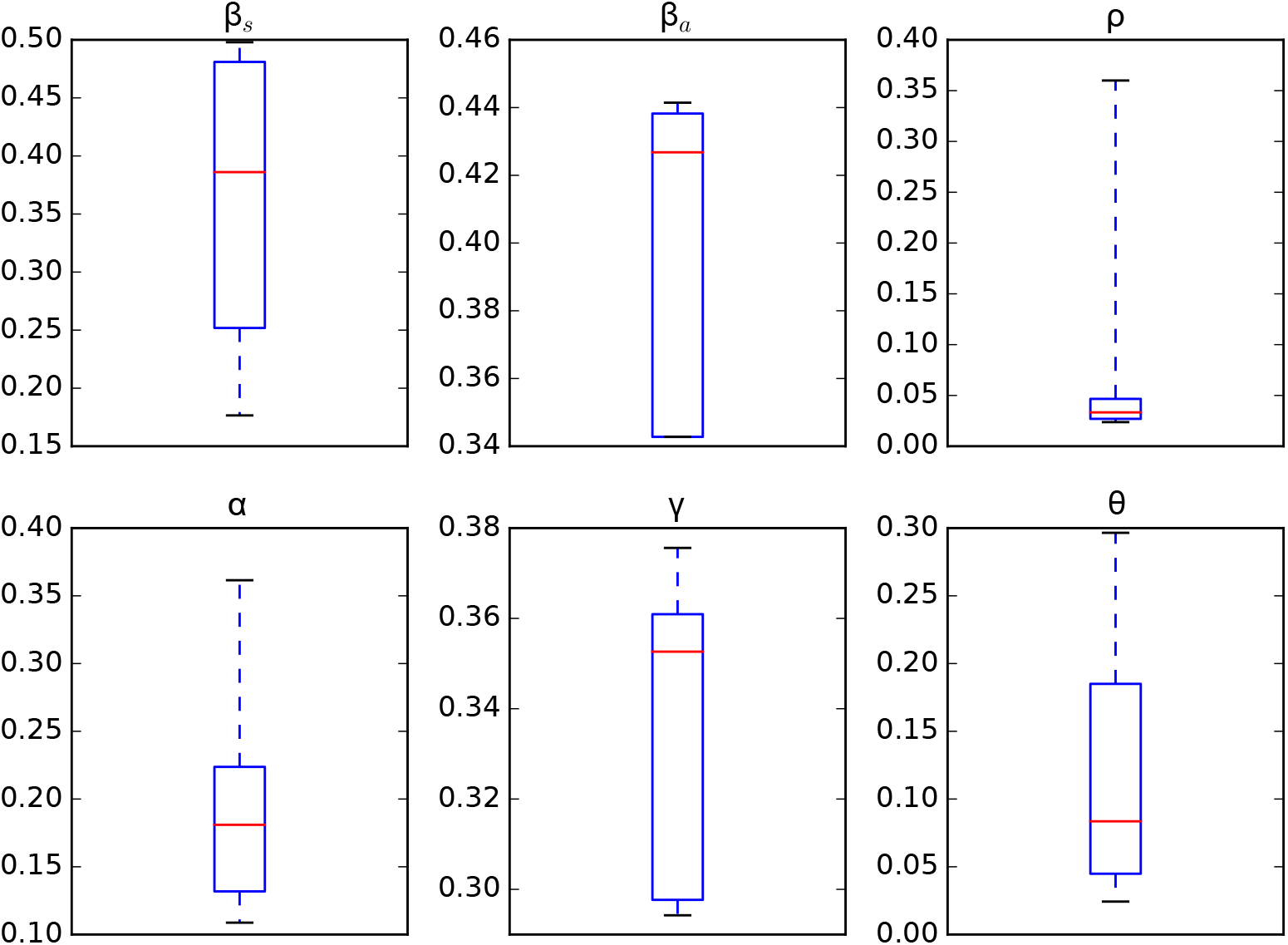
Credible intervals of parameters of model (1) within 95% Highest-Posterior Density (HPD) using the t-walk package [19]

## 6 Discussion

In this work, we analyse a networked dynamic metapopulation model of the coronavirus dissemination in Mexico City using ODEs and Bayesian Statistics. We present an explanation of how to estimate the mobility between the boroughs that compound the Mexico City area both on a weekday and on weekends; we combine information available from the origin-destination survey carried out in 2017 with the current mobility indices that Google and the government of Mexico City report depending on the mode of transport used to make each trip (bus, subway, car, etc.). Also, we present a clustering analysis of the boroughs which compound Mexico City based on mobility data from Google and a Transportation Mexican Survey. From Figure 3, we may identify three different clusters during the each phase of the pandemic. Also, from Figure 4, we may identify four clusters during the four phases of the pandemic. We consider this clustering analysis based on individuals movement may be crucial to model efficiently a human pandemic.

From Figure 5, the transmission rate of symptomatic was 0.19 within 95% Credible Interval (CI) [0.06, 0.42], and the transmission rate of asymptomatic was 0.27 within 95% CI [0.14, 0.40], the fraction of undocumented infections, *ρ*, was 0.027 within 95% CI [0.02, 0.04]. The latency and recovery periods, 1*/α* and 1*/γ*, were 5.96 days within 95% CI [3.60, 8.93] days, and 4.86 days within 95% CI [3.15, 9.33] days respectively. The inter-borough scale factor *θ* was 0.26 within 95% CI [0.15, 0.40], this value indicate that the mean number of trips done by a person is between 3 and 4 in one day, which makes sense with complete trips to get out of home, do some activities (work, shopping, services), and return home. These results of the inferred parameters of model (1) and the population size of boroughs, such as, Iztapalapa and Gustavo A. Madero, explain the fast dispersion of COVID-19 and indicate the challenge of finding strategies to contain it. As mentioned in Section 5, we will analyse the identifiability of the parameters of model (1), that is, the *ρ* parameter since this parameter is multiplied by the period of incubation of the disease, *α*, thus, estimating both parameters simultaneously may lead to non-identifiability difficulty. We may observe this non-identifiability in figures 5, 10 and 11, i.e., different combinations of the model parameters lead to the same “energy” value of the system 1. We point out that the parameters, *β*_*s*_, *β*_*a*_ of the model (1) are considered as global, i.e., they are assumed the same for all the boroughs of Mexico City and all the transportation modes. We will explore in a near future a more robust model which will consider not only local parameters of transmission *β*_*s*_, *β*_*a*_, instead of globally, i.e., a pair of transmission rates *β*_*s*_, *βa* for each borough, but also the interstate and international mobility from/to Mexico City. We will take into account imported cases from the rest of the 30 states of the Mexico country. Also, consider the imported cases from overseas coming by airplane passengers. In addition, we will do a global and local sensitivity analysis of model (1). Also, investigate a spatio-temporal model based on a diffusion partial differential equation model combined with individual movement trends.

## Supporting information

Video containing the diagnosed cases with respect the boroughs

Mobility data

## Data Availability

All the data used was obtainted in the following websites:
https://datos.cdmx.gob.mx/explore/dataset/afluencia-preliminar-en-transporte-publico/table/
https://coronavirus.gob.mx/datos/
https://www.google.com/covid19/mobility/
https://www.inegi.org.mx/programas/eod/2017/
https://www.inegi.org.mx/temas/estructura/default.html#Publicaciones/

https://datos.cdmx.gob.mx/explore/dataset/afluencia-preliminar-en-transporte-publico/table/

https://coronavirus.gob.mx/datos/

https://www.google.com/covid19/mobility/

https://www.inegi.org.mx/programas/eod/2017/

https://www.inegi.org.mx/temas/estructura/default.html#Publicaciones/

## Acknowledgments

K. Prieto wishes to acknowledge the financial support from CONACYT through its program “Cátedras CONACYT”. M.V. Chávez-Hernández wishes to acknowledge the grant FC-2016/1948 from CONACYT, 283 Mexico. Jhoana Romero thanks the support of Fundación Ceiba, Colombia.

## Notes

### Competing Interest Statement

The authors have declared no competing interest.

### Funding Statement

No funding was received

### Author Declarations

We have not used any clinical data

